# Viral load of SARS-CoV-2 across patients and compared to other respiratory viruses

**DOI:** 10.1101/2020.07.15.20154518

**Authors:** Damien Jacot, Gilbert Greub, Katia Jaton, Onya Opota

## Abstract

RT-PCRs to detect SARS-CoV-2 RNA is key to manage the COVID-19 pandemic. We analyzed SARS-CoV-2 viral loads from 22’323 RT-PCR results according to samples types, gender, age, and health units. Viral load did not show any difference across age and appears to be a poor predictor of disease outcome. SARS-CoV-2 viral load showed similar high viral loads than the one observed for RSV and influenza B. The importance of viral load to predict contagiousness and to assess disease progression is discussed.

## 1. Introduction

At the beginning of January 2020, the cluster of SARS-CoV-2 cases identified in Wuhan City, Hubei Province (China) rapidly spread to other regions in China and to other countries, causing a world pandemic (1, 2). Quantitative reverse transcription polymerase chain reaction (RT-PCR) represents a key diagnostic tool for patients with suspected SARS-CoV-2 infection. Viral-specific genes, such as the Envelope (E), the RdRP/Helicase (Hel), the spike protein encoding gene (S), as well as Nucleocapsid (N) were used as molecular targets and combination of these genes have been recommended by the WHO (3, 4). We introduced the E, RdRP, and N genes RT-PCRs in our fully automated molecular diagnostic platform (MDx platform) (5). A lower sensitivity of the RT-PCRS targeting the RdRP and N genes, compared to that targeting the E gene was observed leading us to use solely the E gene, as RT-PCR target. Latter during the pandemic, the cobas® SARS-CoV-2 test (Roche, Basel, Switzerland) became available targeting the ORF1/a, a non-structural region for specific detection of SARS-CoV-2 and a conserved region in the E gene, for pan-Sarbecovirus detection. The pan-Sarbecovirus primers and probe can also detect the SARS-CoV-1 virus, however not currently circulating (6).

We determined the correlation between the cycle threshold (Ct) value and viral load and investigated the distribution of viral loads across sex, age, and healthcare departments and as well as against other respiratory viruses. The report of RT-PCR SARS-CoV-2 viral loads raised also several questions regarding the use of this information for the laboratory as an internal quality assessment tool, as well as (i) to predict contagiousness of patients and hence to guide epidemiological decisions, especially for hospitalized patients and (ii) to predict the patient prognosis and assess disease progression. These important questions will also be discussed here.

## 2. Material and methods

### Data

Data from 19’832 SARS-CoV-2 RT-PCR results from patients with suspected COVID-19 were collected from 1^st^ February to 27^th^ April 2020 at the diagnostic microbiology laboratory of the Lausanne’s University Hospital (CHUV), representing 4172 positive cases.

### SARS-CoV-2 RT-PCR, cycle thresholds and viral load quantification

SARS-CoV-2 was detected in clinical specimen with i) a in-house RT-PCRs targeting the E-gene introduced in our automated molecular diagnostic platform (MDx platform) (5) and with the cobas SARS-CoV-2 test on the cobas 6800 instrument (Roche, Basel, Switzerland). The primers and probes for the E-gene PCR was those described by Corman and colleague (4). Cts of the MDx platform targeting the E gene were converted to viral load using either a plasmid containing the target sequence of the PCR obtained from RD-Biotech (Besançon, France) or using purified viral RNA, kindly provided by the Institute of Virology of the University of Berlin, la Charité (4). Both approaches showed similar virus quantifications and the following equation derived from RNA quantification was used: −0.27Ct+13.04. A comparative analysis of the Cts values obtained from our MDx platform compared to the cobas^®^ SARS-CoV-2 test (Roche, Basel, Switzerland) showed a good congruency for Cts related to the E gene. This led us to use the E gene RT-PCR Cts values of both platforms in the present analyses.

### Clinical specimens

Among the 22’323 specimens collected, only the initial sample per patient was kept (19’832 samples with 4172 positives) and only nasopharyngeal and/or nasal swabs (NPS) were used (19’728 samples). Viral loads in different specimen types were instigated using multiple samples per patient as most of these investigations were performed after the first positive tests, usually an NPS. Comparison of viral load in different hospital unit was also performed using more than one sample per patient.

### Comparison of SARS-CoV-2 with other respiratory viruses

6’050 RT-PCR of 14 other respiratory viruses were extracted from our database over a period of 5 years (2015-2020): Influenza A and B, Respiratory Syncytial Virus (RSV), adenovirus, parainfluenza 1-4, Coronaviruses E229, OC43, HKU1, NL63, Pan-entero/rhinovirus and Human Metapneumovirus and Ct values were obtained on the MDx platform and converted to viral loads, as previously reported (5). For Influenza A and B and RSV the Xpert^®^ Xpress Flu/RSV was used and converted to viral load according to Zou et al. (7). Only nasopharyngeal and nose swabs were included.

### Statistical analysis

Data were process with Rstudio and plotted using ggplot2. Median is presented in all graphs. Statistical significance of viral loads were assessed using a parametric paired t-test and the twotailed p-values interpretation are written on the graphs.

## 3. Results

### SARS-CoV-2 viral load across the pandemic and among other respiratory viruses

We observed a broad distribution of viral load values (Fig. 1A) with an evolution over the pandemic period that mirrored the epidemiological observations of SARS-CoV-2 infection in Switzerland (8) (Fig. 1B). The first cases occurred early March with a peak of the COVID-19 epidemic mid-March followed by a 2 weeks stationary phase before a slow decrease. Interestingly, the median viral load was higher in the first phase of the outbreak as compared to the following period. This is likely linked to the diagnostic of newly infected symptomatic persons with high viral load during the first phase compared to a more heterogeneous population tested in the following months. The initial viral load of SARS-CoV-2 was compared to 14 other respiratory viruses (Fig. 2A) (9). We found that although significant differences in viral loads exist across the different viruses and compared to SARS-CoV-2, SARS-CoV-2 exhibits similar viral load than RSV and Influenza B and than other coronaviruses. The range of viral load is overall similar between all the different respiratory viruses, with some subjects exhibiting very high load while others may exhibit much lower viral load, reflecting likely different sampling times during the course of the disease.

**Figure 1.**
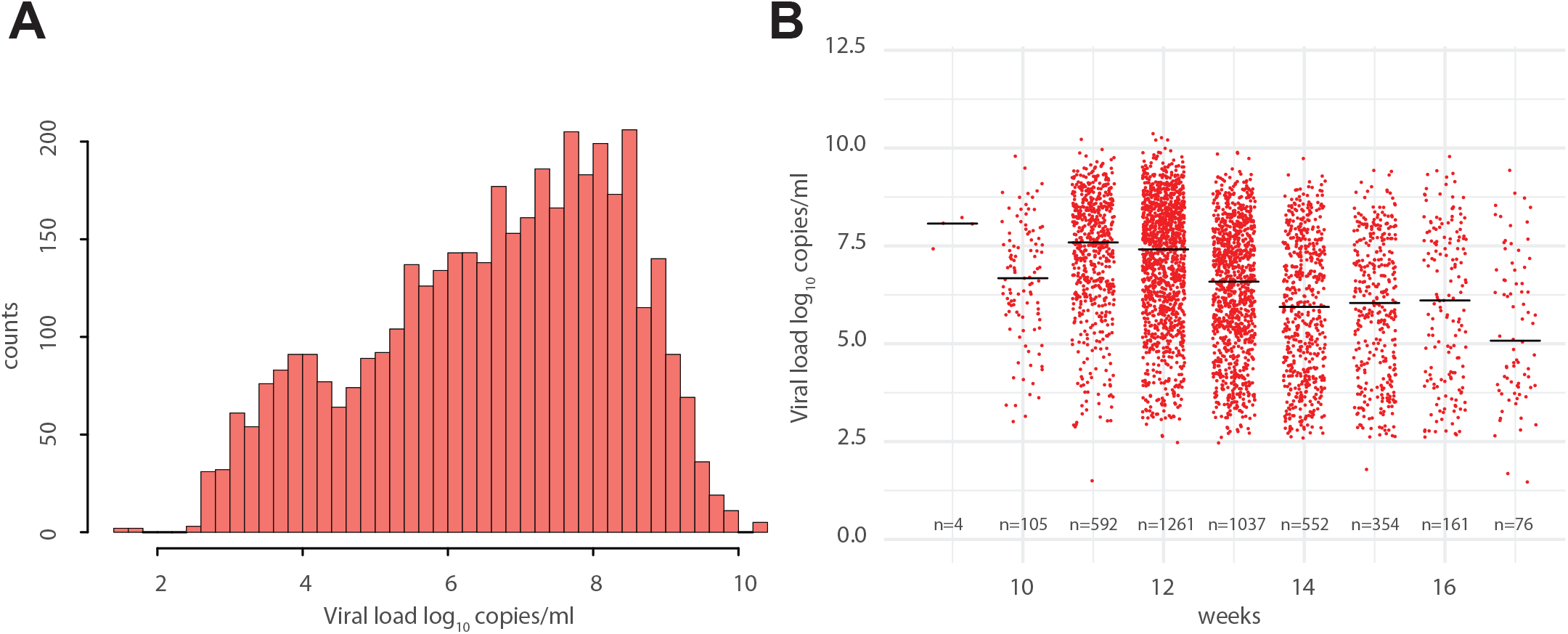
**A:** Histogram of SARS-CoV-2 viral loads. **B:** Time-course analyses of SARS-CoV-2 viral loads across time. Viral loads mirrored the reported COVID-19 infections in Switzerland.

**Figure 2.**
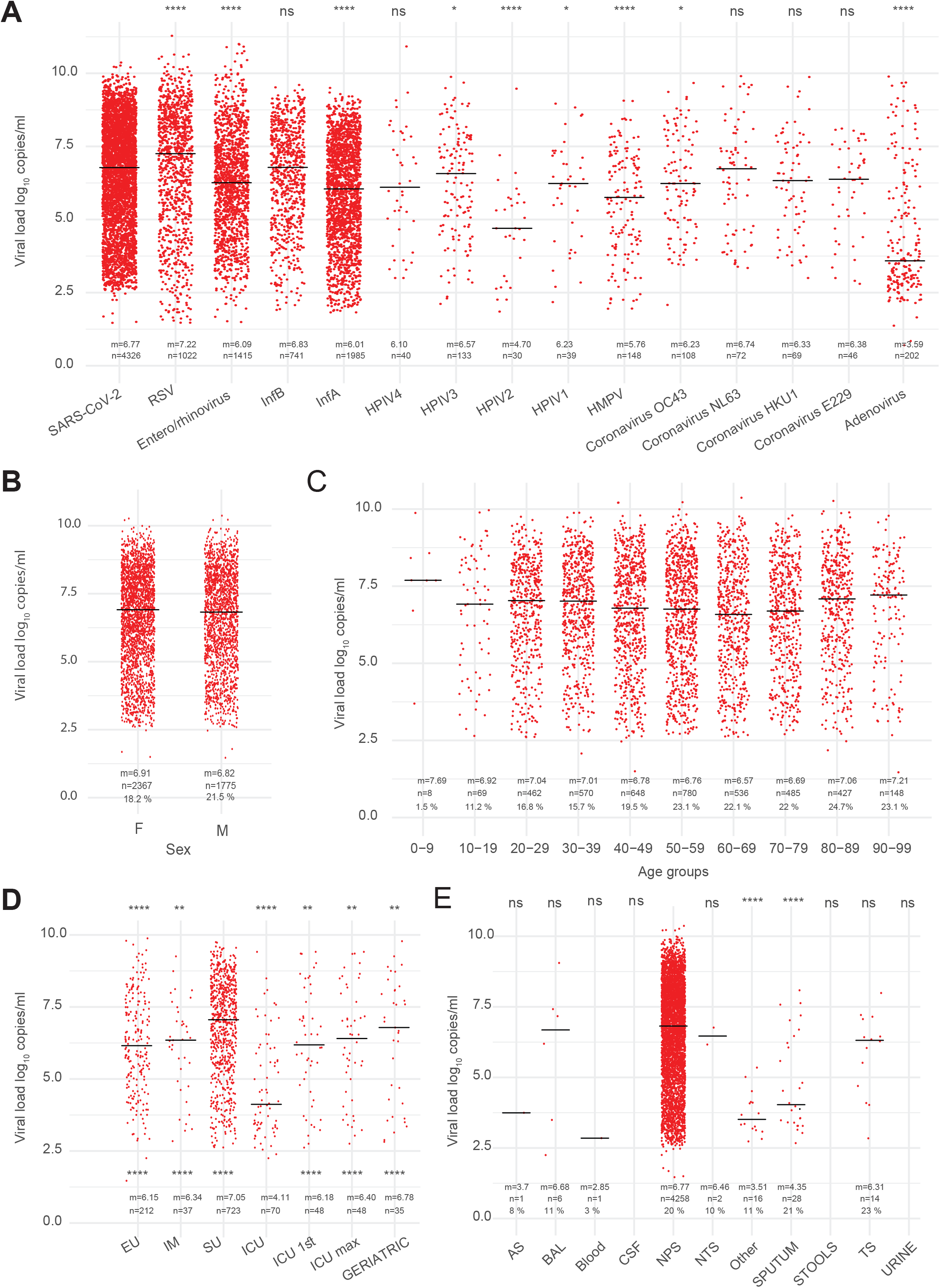
**A:** Viral loads of 14 respiratory viruses compared to SARS-CoV-2. HPMV: Human-metapneumovirus, HPIV1-4: Human Parainfluenza Viruses 1-4, InfA and B: Influenza viruses A and B; RSV: Respiratory Syncytial Virus, m represents the median, n the number of observations, and the percentage of positive test is presented. Statistical significance of viral loads of SARS-CoV-2 was assessed against the other viruses. **B-C:** Viral loads distribution of SARS-CoV-2 across sex and age showed comparable values among all groups. **D:** Initial viral loads of SARS-CoV-2 in different hospital departments. ICU first and ICU max correspond to respectively the first or highest sample recorder for patients latter admitted to the ICU. Statistical significance of viral loads was assessed against the SU samples (upper stars), and against the ICU (lower stars). **E:** Distribution of viral loads across different specimens. Statistical significance of viral loads was assessed against the NPS samples AS: anal swab, BAL: bronchoalveolar lavage, CSF cerebrospinal fluid, NTS: nasal-throat swab, TS: throat swab. P-values: ns: p > 0.05, *: p ≤ 0.05, **; p ≤ 0.01, ***; p ≤ 0.001, ****: p ≤ 0.0001

### SARS-Cov-2 viral loads stratified by gender and age

A higher number of tests was achieved in women than in men (35% of difference); however the rate of positive results was similar for both sex (Fig. S1A and B) and both genders showed comparable viral load distribution (Fig. 2B). Stratification of positive samples by age groups showed that older individuals, when tested, were likely to be proportionally more frequently positive than the rest of the population, while young children showed very low percentages of positivity despite being rarely tested (Fig. S1C-D). Interestingly, viral loads categorization based on 5-year brackets ages showed no significant differences across age groups (Fig. 2C). Although limited by the low samples size, the pediatric age groups showed viral loads values comparable to adults.

### SARS-Cov-2 viral loads across different hospital units

We focused on the Intensive care unit (ICU), the internal medicine (IM) department, the emergency unit (EU) and patients addressed to a screening unit (SU) specifically developed during the outbreak. This stratification per unit was used to investigate possible differences in viral loads in patients with several days of evolution since first symptoms and with a severe lung disease (ICU), versus subjects sick enough to get hospitalized (IM), to patients screened with mild symptoms (SU). To assess if the initial viral load could correlate with disease progression, we traced back, when available, the initial or the highest viral load values obtained in other departments for all patient hospitalized in ICU and showed that this value is not significantly higher than the one obtain for all other patients (Fig. 2D). Interestingly, patients latter hospitalized in ICU showed the lowest viral load in the upper respiratory tract compared to all other patients (Fig. 2D). This might reflect the evolution of COVID-19 infection, from the upper respiratory tract where it causes mild symptoms such as a fever and cough to a more severe form when the lower respiratory tract are affected (10-12). Furthermore, in the secondary phase of the disease, inflammation rather than viral replication appears to predominate (although this was not formerly established in the present work). These observations might also be biased by the timing of the 1^st^ nasopharyngeal test that was sometimes done very late, i.e. at time of admission at the ICU. Finally, geriatric patients did not show different viral loads than other departments.

### SARS-Cov-2 viral loads across different specimens

Over the time course of the epidemic several, non-nasal specimens were analysed mainly lower respiratory samples for patient in the ICU (Fig. S1F). Although, lower viral load values were obtained compared to the upper respiratory part (Fig. 2E), the lower respiratory tract samples were often useful to allow an early microbial diagnostic of COVID-19, and might prove to be useful to assess the clinical prognosis and disease progression. Only few blood samples were tested and only one of them was positive; this suggest a low rate of viremia. Interestingly, SARS-CoV-2 was not detected in urines. This was expected since respiratory tract viruses, which are not associated with a sustained viremia, are unlikely to be shed in urines. Moreover, the absence of virus in the CSF tested samples suggests that the serology should be considered as first line test for meningoencephalitis and Guillain-Barre syndrome. Only a handful number of samples were positive for stools and rectal swabs, due to limited number of subjects tested. Statistical comparison across the different specimens was however limited by the low number of data.

## 4. Discussion

Initial SARS-CoV-2 viral load is widely distributed ranging from 3 to 10 log copies/ml and the evolution of the viral load over-time mirrored the evolution of SARS-CoV-2 infections in Switzerland. The median viral load for SARS-CoV-2 in NPS was 6.78 log_10_ copies per ml. This supports the fact that RT-PCR, which can detect less than 100 copies per ml of samples, is a sensitive method for the diagnostic of COVID-19. This is however limited by the quality of specimen sampling and the time course of infection.

We also compared SARS-CoV-2 viral loads to that of other respiratory viruses in order to determine whether higher viral loads, that could affect contagiousness, are observed. Although significant differences were observed when compared to some other respiratory viruses, SARS-CoV-2 appears to exhibit similar viral load than RSV and influenza B, as previously reported (9). For respiratory viruses other than Influenza and RSV, we have a bias towards immunocompromised or severely ill patients, which might tend to have higher viral loads. Interestingly, others reported that the pattern of patients infected with SARS-CoV-2 resembles more to patients infected with influenza (13) than SARS-CoV-1 (14); the former being characterized by increased infectiousness at time or even before symptoms onset (15). SARS-CoV-2 viral load appears to be a poor predictor of disease outcome. Indeed neither the initial nor the highest viral load of patients latter admitted to the ICU was significantly higher than the specimens from patient treated in a SU. This absence of correlation with the clinical outcome is also supported (i) by other published data showing high viral load in asymptomatic patients (15-18) and (ii) by the fact that asymptomatic or minimally symptomatic patients can transmit the virus (19). We also observed that viral load seems not to correlate with age. In particular, older individual and young children showed similar viral loads than the general population (20-22). Concentration of the virus in the respiratory tract can indirectly reflects contagiousness; however, viral load is not the only factor at play in term of contagiousness, since nasal discharge and cough are clearly important co-variables impacting transmission (23).

The clinical relevance and usefulness of viral load measures appears to be mainly restricted to specifically classifying the patient as being in the first phase of the disease with high viral load or rather in the 2^nd^ phase of the disease when viral load tends to decrease and when inflammation predominates (12). This may be useful to help treatment decision, i.e. to use for instance anti-IL6 or steroids in presence a cytokine storm or during a macrophage activation syndrome. Indeed, COVID-19 disease severity is not directly linked to viral replication in the upper and lower respiratory tracts but is also to an unregulated inflammatory process induced by the host immune response (12). Interpretation of a unique viral load value in a given patients should be done cautiously since (i) there is a trend to a natural gradual decrease of the viral load in the nasopharyngeal samples over time during the course of the infection (15, 16) and (ii) the absolute value of the viral load in the nasopharyngeal samples may be highly different according to the quality of sampling. Despite these limitations, our laboratory decided to provide quantitative results to clinicians, and these values are 172 now used not only for patient care, but also to define contagiousness, i.e. values below 1000 copies/ml may be considered at low risk of transmission. Of course, decisions about patients isolation inside the hospital is not only based on viral load but also takes into account (i) epidemiological aspects such as the possible exposure of other immunocompromised subjects and (ii) clinical presentation, since a patient with cough and/or nasal discharge will be more contagious.

## Data Availability

The readers can contact the corresponding author regarding the data and code discussed in this paper.

## Acknowledgments

We would like to thank all the staff of the Institute of Microbiology of the Lausanne University Hospital. In particular all the biomedical technicians of the molecular diagnostic laboratory for their incredible work and support during the pandemic.

## Conflict of interest

The authors declare to have no conflict of interest.

## Supplementary figure

**Figure 1** A-D: Absolute and percentage values of SARS-CoV-2 infection across sex and ages. E-F: Absolute and percentage values of SARS-CoV-2 viral loads across different specimens. AS: anal swab, BAL: bronchoalveolar lavage, CSF cerebrospinal fluid, NTS: nasal-throat swab, TS: throat swab.

## Notes

### Competing Interest Statement

The authors have declared no competing interest.

### Funding Statement

No external funding was received.

### Author Declarations

The data were obtained during a quality enhancement project at our institution. According to national law the performance and publishing the results of such a project can be done without asking the permission of the competent research ethics committee; exemption letter provided by the "Commission cantonale d'ethique de la recherche sur l'etre humain" of the Vaud Canton (CER-VD), Lausanne, Switzerland.

